# Development and Validation of a Short Form of Suboptimal Health Status Questionnaire

**DOI:** 10.1101/2023.05.24.23290450

**Authors:** Shuyu Sun, Hongzhi Liu, Zheng Guo, Qihua Guan, Yinghao Wang, Jie Wang, Yan Qi, Yuxiang Yan, Youxin Wang, Jun Wen, Haifeng Hou

**Author notes:** **Corresponding to: Haifeng Hou, PhD**, Professor. These authors contributed equally: Shuyu Sun, Hongzhi Liu, Zheng Guo.

## Abstract

**Background:** Suboptimal health status (SHS) is a reversible borderline condition between optimal health and diseases. Although the definition of SHS is widely understood, the questionnaires of SHS are needed to be further developed, by which individual with SHS can be identified from a variety of population in the context of predictive, preventive and personalized medicine (PPPM/3PM). This study aimed to develop a short form of suboptimal health status questionnaire (SHSQ-SF) by reference to suboptimal health status questionnaire-25 (SHSQ-25).

**Methods:** A total of 6,183 participants enrolled from southern China were included in a training set, while 4,113 from northern China were included in an external validation set. SHSQ-SF included nine key items from SHSQ-25, a questionnaire that has been applied in Caucasians, Asians, and Africans. Item analysis, reliability and validity tests were carried out to validate SHSQ-SF. Receiver operating characteristic (ROC) curve was used to identify the optimal cutoff value for diagnosis of SHS.

**Results:** In the training dataset, the Cronbach’s α coefficient was 0.902, and the split-half reliability was 0.863. The Kaiser-Meyer-Olkin (KMO) statistic was 0.880, and the Bartlett’s test of sphericity was significant (χ*^2^* = 32,929.680, *P<*0.05). Both Kaiser’s criteria (eigenvalues >1) and scree plot revealed one factor which explained 57.008% of the total variance. Standardized factor loadings of confirmatory factor analysis (CFA) indices were between 0.59 to 0.74, with χ*^2^*/*dƒ* = 4.972, (GFI) = 0.996, CFI = 0.996, RFI = 0.989 and RMSEA = 0.031. The area under ROC curve (AUC) was 0.985 (95%CI: 0.983 – 0.988) in training dataset, by which the cutoff value (≥ 11) was identified for diagnosis of SHS. In the external validation dataset, this questionnaire showed good discriminatory power (AUC = 0.975, 95%CI: 0.971 – 0.979), with a sensitivity of 96.2% and specificity of 87.4%.

**Conclusions:** We developed a short form of SHS questionnaire, which has good reliability and validity in measurement of SHS in Chinese residents. From the perspective of PPPM/3PM, SHSQ-SF is recommended to be used for quickly screening individuals with SHS from a large-scale population.

## Introduction

The World Health Organization (WHO) defines optimal health as “a state of complete physical, mental and social well-being and not merely the absence of disease or infirmity” [1]. Although the health care system has played a key role in control of chronic disease, there are an increasing number of individuals who suffer suboptimal health status (SHS) in China, a condition between optimal health and a diagnosable disease [2]. SHS are characterized by the perception of health complaints, chronic fatigue, and a constellation of physical symptoms such as the cardiovascular system, the digestive system, the immune system, and mental status; lasting for at least 3 months [2, 3]. In a global survey, the WHO released that 75% of the people are at the risk of suboptimal health, and only 5% are of the real healthy people in the world [4]. From 2007 to 2011, a national survey identified that the prevalence of SHS was 69.46% across China [5]. A variety of studies focusing on SHS in different occupations and ages revealed that the prevalence rates of SHS range from 18.67% to 50.70% [6–10], posing a major global public health challenge in China [11, 12].

Although the causes of SHS are unclear, it is believed that SHS shares similar behavior-related risk factors with non-communicable diseases (NCDs), such as physical inactivity, poor diet, psychological stress, smoking, and excessive alcohol consumption [13, 14]. SHS induces unbalance or symptoms in cardiovascular [11, 15], digestive [16], immune [17] and mental systems [18]. If these symptoms cannot be relieved or reversed, SHS has high likelihood in leading to irreversible pathology and associated complications within a few years [19]. Previous studies have presented that SHS might represent the period preceding the occurrence of clinical manifestations of NCDs, such as cardiovascular disease (CVD) and diabetes mellitus type 2 (T2DM) [20, 21]. From the standpoint of predictive, preventive and personalized medicine (PPPM/3PM) that a new paradigm of public health services technique intensively expanded worldwide, the concept of SHS reflects the viewpoint NCDs can be effectively predicted and prevented before the occurrence of a clinical manifestation of severe pathologies, thus early identification of SHS creates a window opportunity in the targeted prevention and personalized treatment of NCDs [3]. PPPM/3PM is a concept enabling the prediction of an individual’s predisposition to disease before its onset, followed by preventive measures delivered via personalized treatment algorithms. Therefore, focusing on SHS, early diagnosis and targeted intervention approaches in terms of PPPM/3PM have been explored to minimize the risk of SHS precede the occurrence of NCDs [11, 22–24]. As such, a series of SHS questionnaires have been established, such as suboptimal health status questionnaire-25 (SHSQ-25), Multidimensional Sub-health Questionnaire of Adolescents (MSQA), Sub-health Measurement Scale Version1.0 (SHMS V1.0), Chinese Sub-health State Evaluation Scale (CSHES), and Sub-Health Self-Rating Scale (SSS) [22, 25–27], among which SHSQ-25 is the most widely used scale globally with the highest quality in the line with PPPM/3PM [28].

In order to conduct an investigation on health status, psychology, behavior, and social determinants in Chinese residents from 148 cities [29], we developed a short-form questionnaire of SHS, by which we proposed to identify the profile of SHS in the nation-wide cross-sectional study.

## Methods

### Study participants

We selected two samples in China *via* a convenient sampling method: (1) persons from Kunming City, Yunnan Province as a training set; (2) population from Taian City, Shandong Province as an external validation set (Fig. 1). The inclusion criteria were: (1) age ≥ 16 years old; (2) no somatic abnormalities or psychiatric abnormalities currently; (3) no history of medication consumption in the previous 2 weeks. The exclusion criteria were: (1) history of somatic diseases; (2) history of psychiatric abnormalities.

**Fig. 1.**
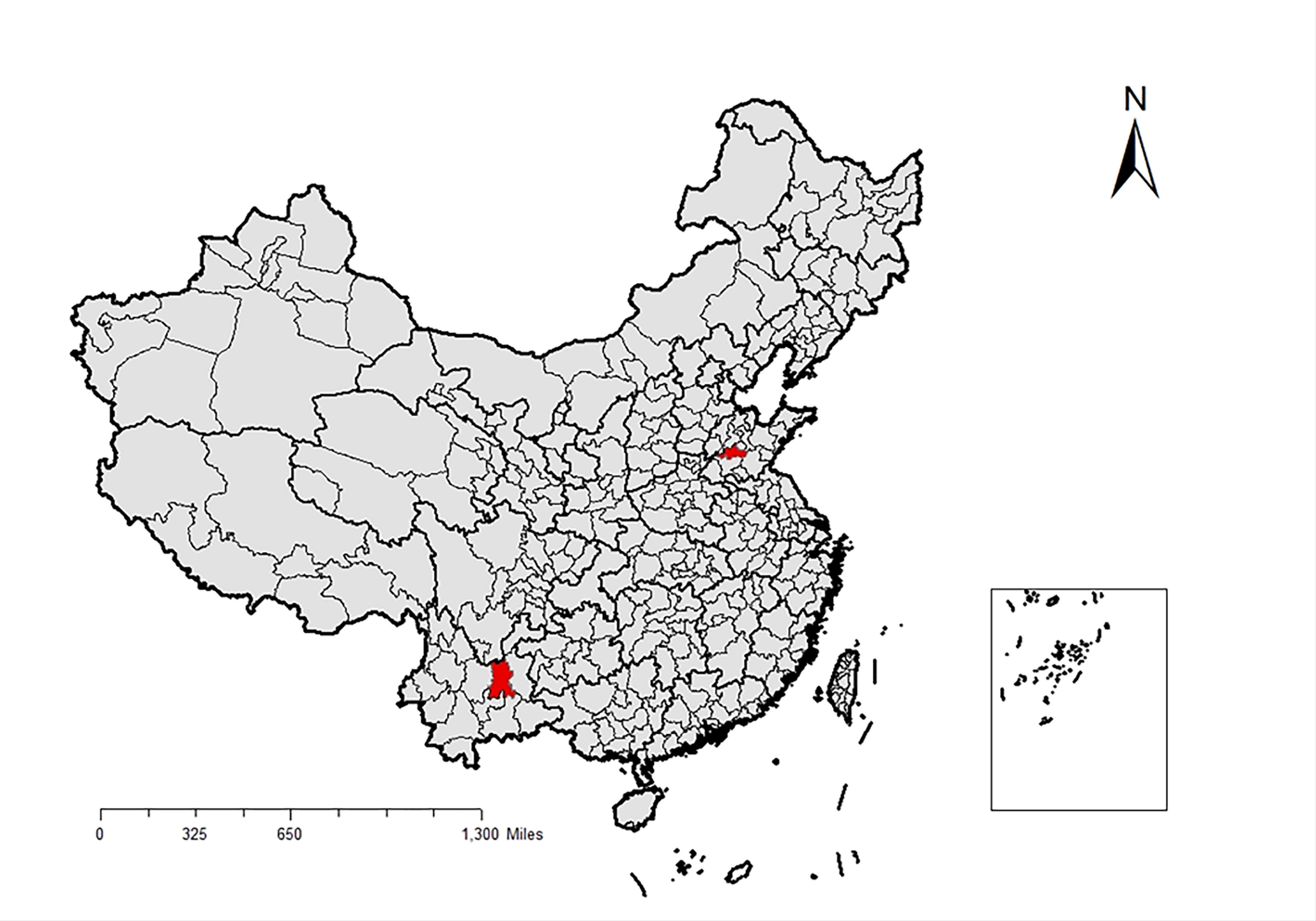
Location of the regions in China

### Research tools

We investigated the demographic information (i.e., gender, age, residence area) behavioral lifestyles and SHS.

#### Measurement of SHS

The health status of participants was evaluated by the SHSQ-25 which includes 25 items in five dimensions: (1) fatigue, (2) cardiovascular health, (3) the digestive tract, (4) immune system and (5) mental health [22]. Each item had 5 options: (1) never or almost never, (2) occasionally, (3) often, (4) very often and (5) always, which were assigned scores of 0-4, respectively[11]. The score of each item was summed together to obtain the total score of SHSQ-25 [22], by which the participants were classified into ideal health (with a summed score <35) and suboptimal health (with a summed score ≥35) groups.

#### Development of short-form suboptimal health status questionnaire (SHSQ-SF)

To determine which items to retain on the brief version of the SHS questionnaire, we used the results of the exploratory factor analysis (EFA) in the Development and Evaluation of a Questionnaire for Measuring Suboptimal Health Status in Urban Chinese [22, 30, 31].

### Statistical analysis

Data with inconsistent logic checks, incomplete information, and data where the options checked were all the same were removed. During development of SHSQ-SF based on the training dataset, we used item analysis, reliability analysis and EFA to test the reliability and validity of SHSQ-SF. Moreover, the confirmatory factor analysis (CFA) and receiver operating characteristic (ROC) curve were performed with the external validation dataset.

The correlation coefficient, extreme group method, and corrected item total correlation (CITC) method were used for item analysis [32]. In the correlation coefficient analysis, the items with coefficients r <0.40 and *P*>0.05 to the total score were deleted. For the extreme group method, each item was compared between the participants with total score at the highest 27% and the lowest 27%, and those with no significant difference were removed. With regard to the CITC method, if the Cronbach’s α coefficient changes significantly after deletion of an item, it means the item is not appropriate, and will be dropped [32, 33].

Cronbach’s α coefficient and split-half reliability were used to test the reliability of the questionnaire, where α ≥0.70 was regarded as adequate internal consistency [34, 35].

EFA was performed to test the structural validity of the questionnaire. The Kaiser-Meyer-Olkin (KMO) value and Bartlett’s test of sphericity were used to measure the appropriateness of conducting an EFA. If the KMO value ≥ 0.60 and Bartlett’s test of sphericity significantly (*P*<0.05), the data are appropriate for conducting an EFA [36]. Factor loading for each item >0.40 and cumulative variance contribution rate >40% were considered as construct validity [37].

CFA was performed to test the structural validity and the convergent validity of the questionnaire. The structural validity was considered well fitted when (1) χ^2^ over degrees of freedom (χ^2^/*dƒ*) <5, (2) goodness-of-fit index (GFI) >0.9, (3) normed fit index (NFI) >0.9, (4) comparative fit index (CFI) >0.9, (5) relative fit index (RFI) >0.9, (6) root mean square error of approximation (RMSEA) <0.05 [38–40]. In addition, average variance extracted (AVE) and composite reliability (CR) were used to test the convergent validity of the questionnaire [41]. The criteria for good convergent validity were AVE > 0.50 and CR>0.70 [42, 43]. Studies also showed that AVE ≥ 0.50 indicates good convergence validity, whenever AVE < 0.50, CR should surpass 0.70 [41].

A ROC curve was generated based on the training set, and the cutoff value was established by the sensitivity (Se), specificity (Sp), and Youden’s index (YI) [44]. In external validation dataset, the performance of SHSQ-SF in discrimination of individuals with SHS was evaluated by the area under the curve (AUC), which ranges from 0.5 (the test being evaluated has no accuracy) to 1.0 (perfect accuracy) [45]. *Kappa* coefficient was used to evaluate the level of agreement between SHSQ-25 and SHSQ-SF.

All data were analyzed with the SPSS 26.0 (IBM, Armonk, NY, United States) and AMOS 26.0 (IBM, NY, United States). All data were tested using a two-sided test, and *P*<0.05 was considered statistically significant.

## Results

### Characteristics of participants

A total of 6,183 participants enrolled from southern China (Yunnan Province) were included in a training set, while 4,113 from northern China (Shandong Province) were included in an external validation set. The characteristics of all participants who met the inclusion and exclusion criteria are provided in Table 1. In the training set, 5,377 (87.0%) were healthy, while 806 (13.0%) were in SHS. In the external validation set, 3,251 (79.0%) were healthy and 862 (21.0%) were in SHS.

**Table 1.** Characteristics of study participants

| Variables |  | Training set (n=6183) | Validation set (n=4113) |
| --- | --- | --- | --- |
| **Age (mean ±SD)** |  | 20.17±1.96 | 18.75±0.88 |
| **Gender (n, %)** | Male | 1446(23.4) | 1111(27.0) |
|  | Female | 4737(76.6) | 3002(73.0) |
| **Smoking (n, %)** | No | 5796(93.7) | – |
|  | Yes | 387(6.3) | – |
| **Alcohol use (n, %)** | No | 5674(91.8) | – |
|  | Yes | 509(8.2) | – |
| **Area (n, %)** | Rural | 2655(42.9) | 2850(69.3) |
|  | Urban | 3528(57.1) | 1263(30.7) |
| **SHS (n, %)** | No | 5377(87.0) | 3251(79.0) |
|  | Yes | 806(13.0) | 862(21.0) |

SD, standard deviation; SHS, suboptimal health status

### Development of SHSQ-SF

To maintain contents matching the original 25-item questionnaire, we retained four items for the dimension of the fatigue, two items for the dimensions of cardiovascular health and mental health, and one item for the dimension of the digestive tract. As the dimension of the immune system explained relatively lower proportion of total variance of SHS, no items were selected from this dimension. Therefore, the final SHSQ-SF consists of nine items: (1) exhausted without physical actives significantly increasing, (2) languid when working, (3) muscles or joints feel stiff, (4) have pains in shoulder/neck/waist, (5) got out of breath while sitting still, (6) suffered from chest congestion, (7) got poor appetite, (8) have trouble with impairment in short memory, and (9) could not respond quickly (Table S1).

### Item analysis

As shown in Table S2, there were significant correlations between each of the nine items and the total score of SHSQ-SF, with correlation coefficients of 0.682 to 0.826 (*P*<0.05). As shown in Table S3, the difference in each item were significant between individuals in the high (with the highest 27% of SHSQ-SF scores) and low (with the lowest 27% of SHSQ-SF scores) subgroups (*P*<0.05). As shown in Table 2, the CITC were all above 0.601, and the deleted Cronbach’s α showed that the internal consistency coefficients did not change significantly after the deletion of each item (Table 2). All the results evidenced that the SHSQ-SF has good discriminatory power.

**Table 2.** Results of CITC in training set (n=6,183)

| **Items** | **Total correlation** | **$\alpha$** |
| --- | --- | --- |
| Exhausted without physical actives significantly increasing | 0.644 | 0.894 |
| Languid when working | 0.630 | 0.894 |
| Muscles or joints feel stiff | 0.694 | 0.889 |
| Have pains in shoulder/neck/waist | 0.674 | 0.891 |
| Got out of breath while sitting still | 0.675 | 0.891 |
| Suffered from chest congestion | 0.667 | 0.893 |
| Got poor appetite | 0.601 | 0.896 |
| Have trouble with impairment in short memory | 0.750 | 0.884 |
| Could not respond quickly | 0.766 | 0.883 |

CITC, corrected item total correlation; $\alpha$ , Cronbach's alpha after deletion

### Reliability analysis of the SHSQ-SF

The Cronbach’s α coefficient of the SHSQ-SF was 0.902, indicating a high reliability. In addition, the split-half reliability was 0.863, which were above 0.70.

### Validity analysis of the SHSQ-SF

#### Structural validity

An EFA was performed to examine the internal factor structure of the SHSQ-SF. The statistic of KMO test was 0.880, and Bartlett’s test of sphericity was significant (χ^2^ = 32,929.680, *P*<0.05), suggesting appropriate for EFA. On the basis of Kaiser’s criteria (eigenvalues >1) and the scree plot, we extracted one factor (Fig. 2), which explained 57.008% of the total variance (Table 3).

**Fig. 2.**
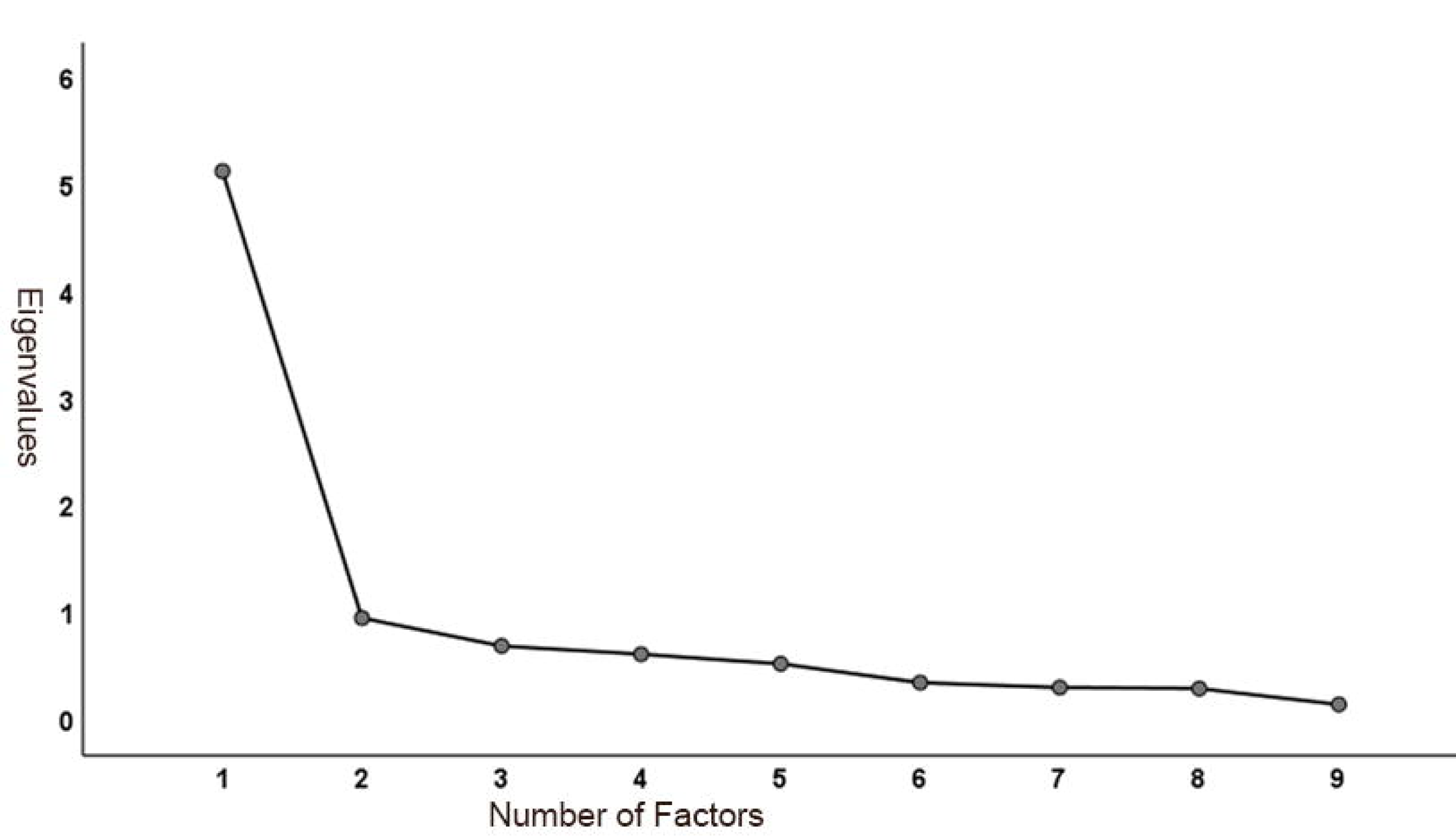
Scree plot for the SHSQ-SF

**Table 3.** Results of EFA with 9 items in training set (n=6,183)

| **Factor/Item** | **Factor loading** | **Eigenvalue** | **Variance explained (%)** |
| --- | --- | --- | --- |
| SHS | – | 5.131 | 57.008 |
| Exhausted without physical<br>activities significantly increasing | 0.711 | – | – |
| Languid when working | 0.695 | – | – |
| Muscles or joints feel stiff | 0.772 | – | – |
| Have pains in<br>shoulder/neck/waist | 0.750 | – | – |
| Got out of breath while sitting<br>still | 0.764 | – | – |
| Suffered from chest congestion | 0.756 | – | – |
| Got poor appetite | 0.691 | – | – |
| Had trouble with impairment in<br>short memory | 0.816 | – | – |
| Could not respond quickly | 0.828 | – | – |

EFA, exploratory factor analysis; SHS, suboptimal health status

By external validation dataset, we performed a CFA to identify the structural validity of the questionnaire. After revised for 12 times *via* the Modified Index (MI), the standardized factor loadings of the CFA were between 0.59 to 0.74 (Fig. 3) (Table 4). And the model fit indexes were as follows: (1) χ^2^/*dƒ* =4.972 <5, (2) GFI=0.996 >0.9, (3) CFI=0.996 >0.9, (4) RFI=0.989 >0.9, (5) NFI=0.995 >0.9 and (6) RMSEA= 0.031 <0.05. These findings indicated that the model structural validity is good and met the requirements.

**Fig. 3.**
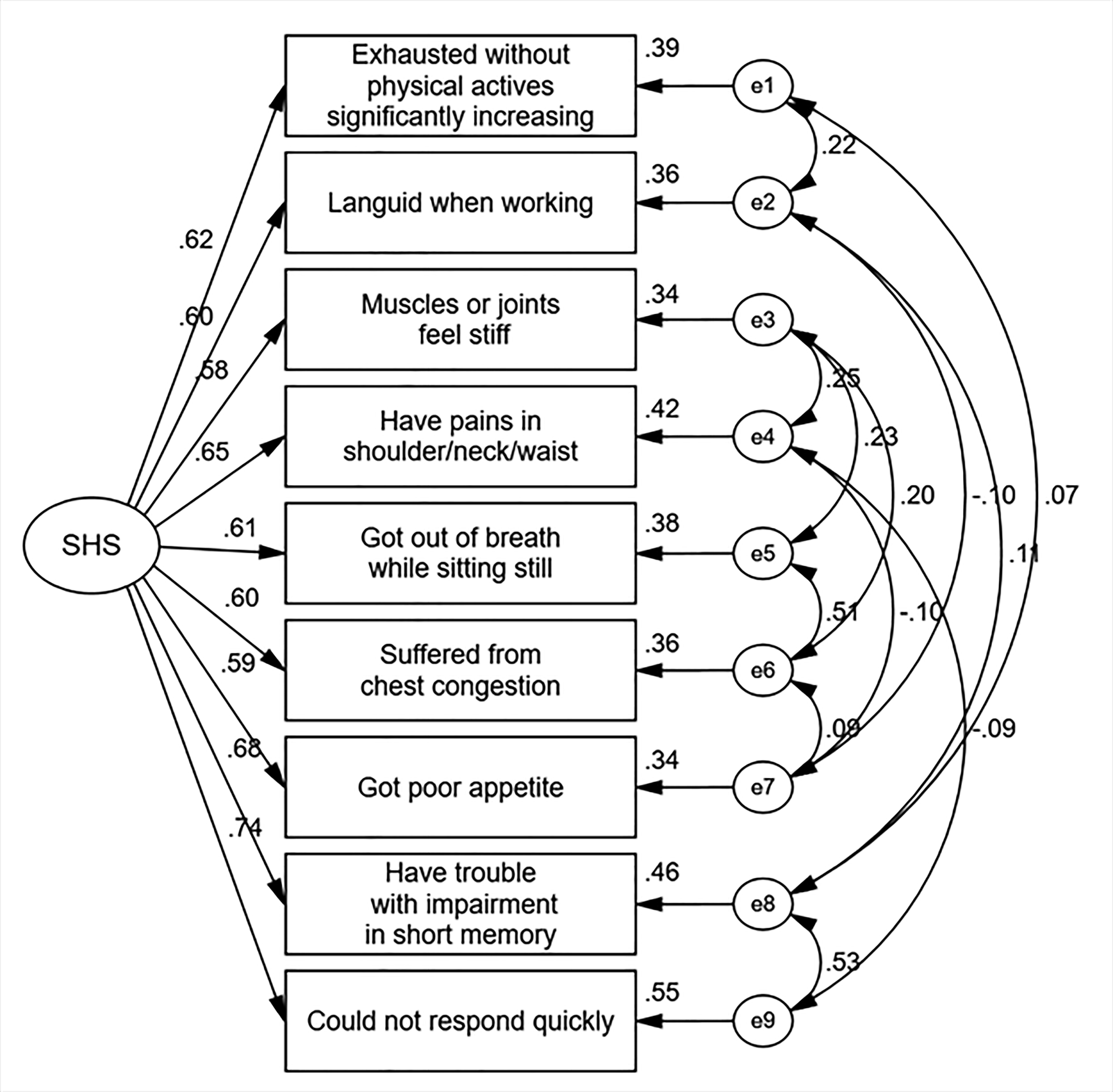
The validation factor analysis model for the SHSQ-SF

**Table 4.**
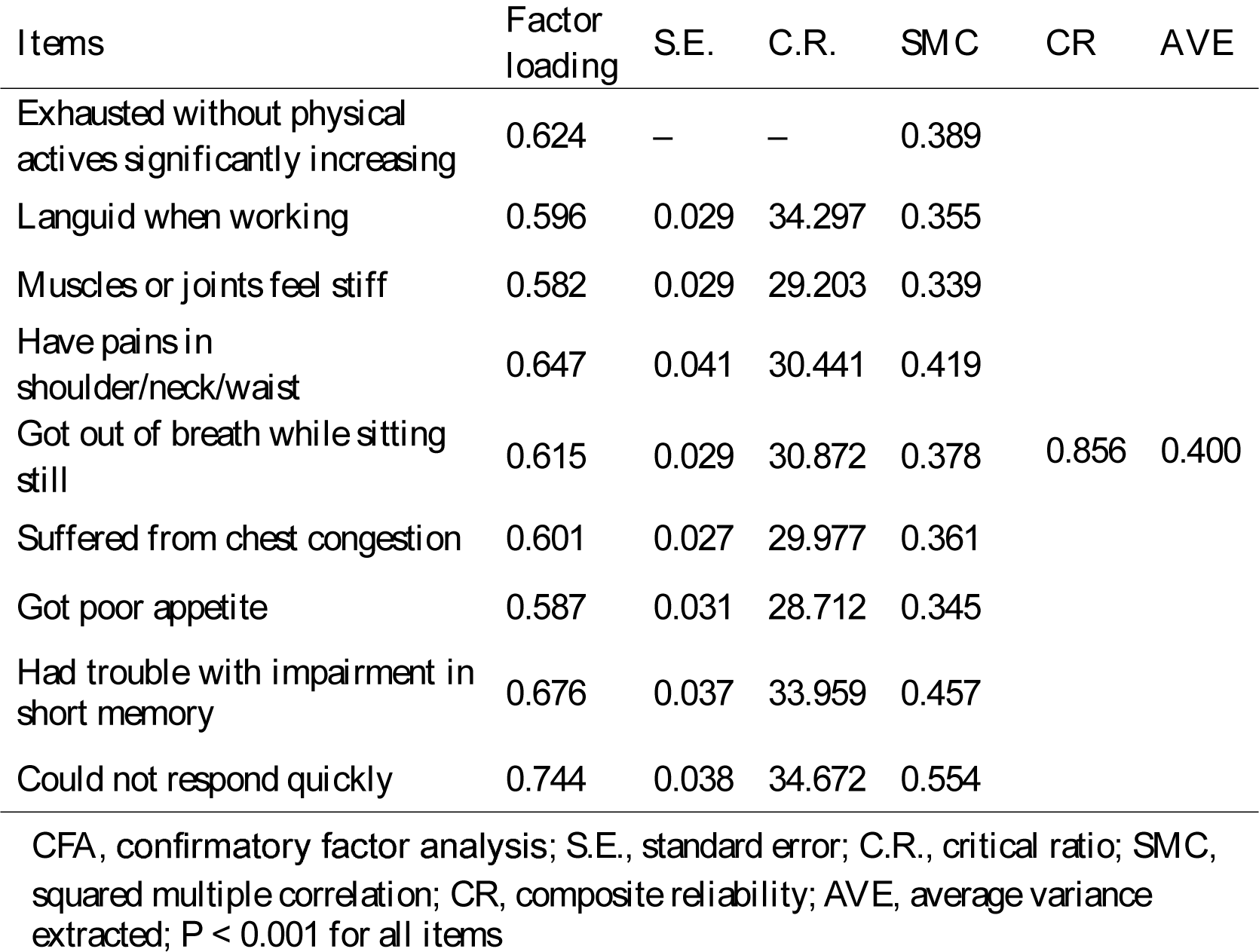
Results of CFA with 9 items in validation set (n=4,113)

#### Convergent validity

The factor had an AVE of 0.400, with the CR of 0.856. This showed that the factor structure had sufficient convergent validity (Table 4).

### The optimal cutoff value and external validation

The AUC was 0.985 (*P*<0.001) in training dataset, which denotes a good diagnostic accuracy of the SHSQ-SF (Fig. 4). By Youden index, the optimal cutoff value (10.5) was selected to identify individuals with SHS, with Se of 96.5% and Sp of 91.5%, respectively (Table S4). Therefore, we used the summed score ≥ 11 as the diagnostic level of SHS. In addition, the *kappa* coefficient between SHSQ-25 and SHSQ-SF was 0.718 (*P*<0.001), indicating strong agreement between the original and simplified questionnaires of SHS.

**Fig. 4.**
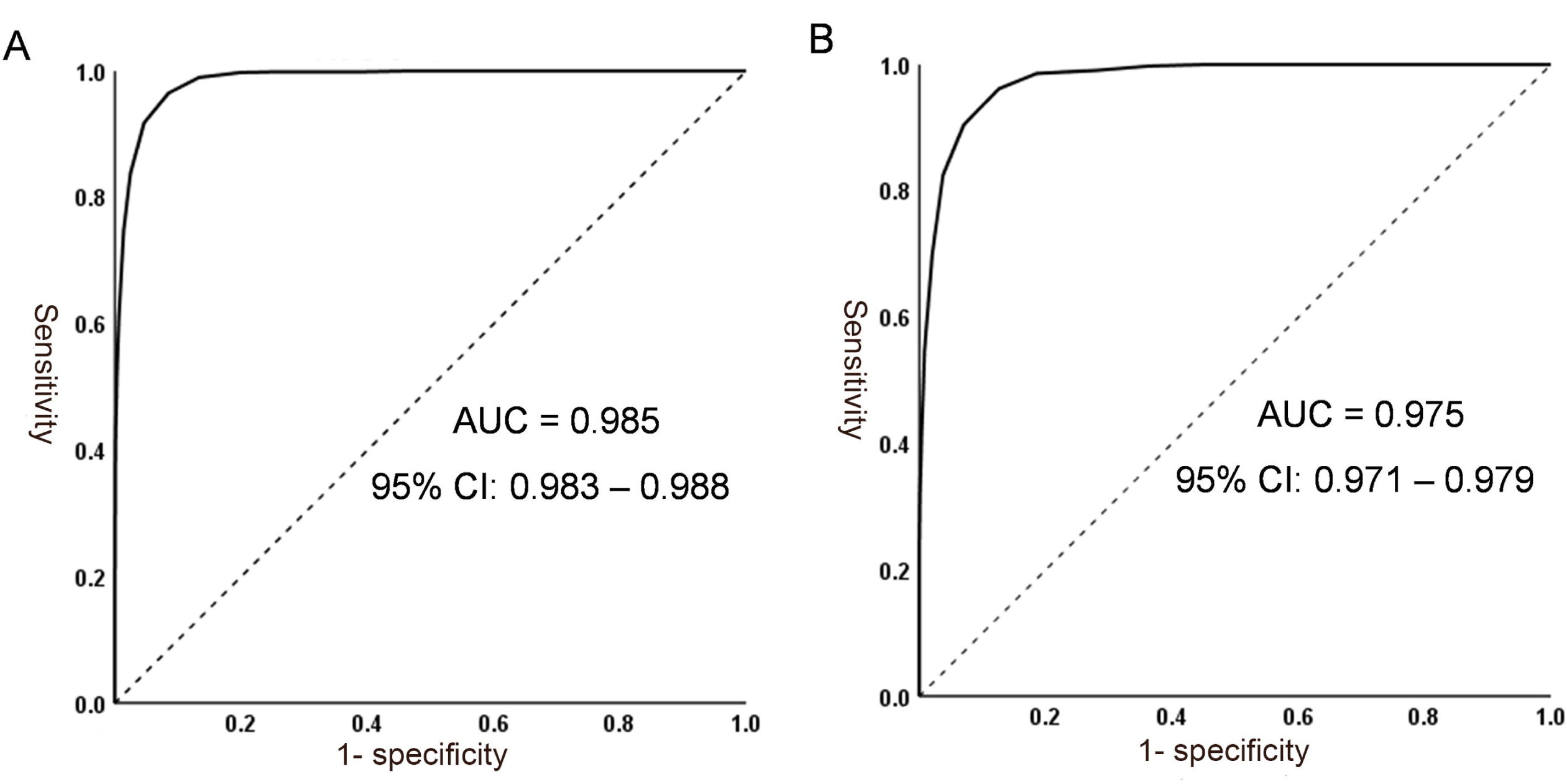
Receiver operating characteristic (ROC) curve. (A) SHSQ-SF for diagnosis of SHS in training set; (B) SHSQ-SF for diagnosis of SHS in validation set

In external validation, SHSQ-SF showed AUC of 0.975 (*P*<0.001), with Se of 96.2%, Sp of 87.4%, and *Kappa* of 0.720 (Fig. 4 and Table S5). These results showed that the high diagnostic performance of SHSQ-SF.

## Discussion

This study developed a short-form questionnaire of SHS (SHSQ-SF), which included nine key items from SHSQ-25. This simplified questionnaire has the highest factor load in the original SHSQ-25 questionnaire, explaining 57.01% of the total variance of SHS [46, 47]. Although SHSQ-SF is a unidimensional measure of SHS, it can evaluate the SHS efficiently when survey space is restricted. This makes it suitable in study with a large-scale population. Except SHSQ-25, there are other SHS scales, such as MSQA, SHMS V1.0, CSHES and SSS [22, 25–27]. These questionnaires contain a very large number of items. SSS consists of three symptom dimensions, with ten factors supported by 58 items, and CSHES include three symptom dimensions, with 18 factors supported by 64 items [26].

SHSQ-SF consists of nine items that represent the healthy complains of the participants in the past three months: (1) exhausted without physical actives significantly increasing, (2) languid when working, (3) muscles or joints feel stiff, (4) have pains in shoulder/neck/waist, (5) got out of breath while sitting still, (6) suffered from chest congestion, (7) got poor appetite, (8) have trouble with impairment in short memory, and (9) could not respond quickly. Each item had 5 options: (1) never or almost never, (2) occasionally, (3) often, (4) very often, and (5) always, which were assigned scores of 0-4, respectively. The score of each item was summed together to obtain the total score of SHSQ-SF, by which the participants were diagnosed with ideal health (with a summed score <11) and suboptimal health (score ≥11). This cutoff point for SHS was established by the ROC curve using the training dataset with an AUC of 0.985. In external validation, SHSQ-SF showed AUC of 0.975, with Se of 94.2%, Sp of 87.4%, and *Kappa* of 0.720. Overall, the SHSQ-SF was highly effective at identifying persons with SHS. The AUC of SHSQ-SF was greater than that of SSS, which indicates this questionnaire was more effective in identifying individuals with SHS. However, other questionnaires did not report such results.

The measurement of each of the nine items of SHS-SF is significantly different between individuals with higher SHS scores (highest 27%) and those with lower scores (lowest 27%), indicating that this scale has potential to discriminate persons at high risk of SHS from the healthy population. Meanwhile, the significant correlation between each item and the summed SHS score (ranging from 0.682 to 0.826), illustrated the homogeneity of this questionnaire [48]. In addition, the sensitivity analysis that was performed with CITC evidenced the stability of the questionnaire.

With regard to reliability and validity, SHSQ-SF also showed consolidating results. In the reliability test, a Cronbach’s α between 0.7 and 0.8 represents fairly good and 0.8 to 0.9 is very good [49]. In our study, the Cronbach’s α of the SHSQ-SF was 0.902, with a split-half reliability of 0.863, suggesting high internal consistency. For the validity test, we examined structural validity and convergent validity. All the modified model fit indices (*i.e.*, χ*^2^/dƒ*, GFI, NFI, RFI, RMSEA) demonstrated the good validity [50, 51]. In terms of convergent validity, although one of statistics (AVE value) was mildly low, the other value (CR) was significantly high instead [52]. This indicated that the factor structure had sufficient convergent validity. Cronbach’s α of SHSQ-SF is consistent with the results of SHSQ-25, MSQA, SHMS V1.0, CSHES and SSS, all of which are greater than 0.8, showing very high internal consistency [22, 25–27].

Since early detection followed by appropriate intervention is important for prevention from the onset of diseases, it should be requested for clinicians to shift from the perspective of delayed intervention approaches to early screening of individuals with SHS. The emergence of PPPM/3PM is the way forward for medical approach as this concept can early identify population who may have SHS and are at risk of developing NCDs [53]. Although the concept of SHS is well understood of an intermediate status between disease and health [54], identification of SHS remains elusive. A full evaluation of health conditions is necessary for the effective implementation of public health intervention [55]. A nation-wide investigation on health status, psychology, behavior, and social determinants has been carried out in China since the year of 2020 [56]. In the framework of PPPM/3PM, the measurement of SHS has been involved in the 2023 proposal of this investigation. We developed this short-form questionnaire of SHS, by which we aimed to identify the profile of SHS in this nation-wide cross-sectional study. To our knowledge, SHSQ-SF is the first questionnaire simplified for measurement of SHS, which is easy to use in large scale population-based studies. The simplified SHSQ-SF, a more cost-effective and powerful SHS assessment tool, would serve to overcome the economic barriers associated with a lack of laboratory tests and treatments, facilitating PPPM/3PM, thus having a significant impact to healthcare.

### Limitations

There are several limitations in this study. First, most of the participants in this study were young people. The old persons who have somatic and psychological disorders were excluded at enrollment. This might limit the representativity of our findings. Second, there is no objective or physical measurement standard for SHS, hence we used SHSQ-25 as the golden standard, which is the widely used scale in Africa, Asia, Europe, and Oceania. Finally, this study may include population who may have disease but were not diagnosed. Therefore, it can be expected that the reported SHS level is probably overestimated.

### Conclusions and expert recommendations

We developed a short form of SHS questionnaire (SHSQ-SF) based on the integrative concept of PPPM/3PM, and SHSQ-SF was validated with good reliability and validity in measurement of SHS in Chinese residents. SHSQ-SF could be recommended to be applied in screening individuals with SHS in a large population as a cost-effective and time-efficient assessment tool in the line with the purpose of PPPM/3PM.

### Declarations

### Abbreviations

SHS: Suboptimal health status; PPPM/3PM: predictive, preventive and personalized medicine; SHSQ-SF: short form of suboptimal health status questionnaire; SHSQ-25: suboptimal health status questionnaire-25; ROC: receiver operating characteristic; KMO: Kaiser-Meyer-Olkin; CFA: confirmatory factor analysis; AUC: area under ROC curve; WHO: World Health Organization; NCDs: non-communicable diseases; CVD: cardiovascular disease; T2DM: diabetes mellitus type 2; MSQA: Multidimensional Sub-health Questionnaire of Adolescents; SHMS V1.0: Sub-health Measurement Scale Version1.0; CSHES: Chinese Sub-health State Evaluation Scale; SSS: Sub-Health Self-Rating Scale; EFA: exploratory factor analysis; CITC: corrected item total correlation; GFI: goodness-of-fit index; NFI: normed fit index; CFI: comparative fit index; RFI: relative fit index; RMSEA: root mean square error of approximation; AVE: average variance extracted; CR: composite reliability; Se: sensitivity; Sp: specificity; YI: Youden’s index; MI: Modified Index; SD: standard deviation; S.E.: standard error; C.R.: critical ratio; SMC: squared multiple correlation.

## Funding

This work was supported by Natural Science Foundation of Shandong Province (ZR2022MH082) and Scientific Research Foundation of Education Department of Yunnan Province (2021J1359).

### Ethics approval

The Ethics Committee of Shandong First Medical University (SDFMU) approved this study.

### Consent to participate

All procedures performed in the study involving human participants were in accordance with the principles outlined in the Helsinki Declaration. All participants were required to sign an informed consent form before being enrolled in this study.

## Consent for publication

All authors have given consent for publication.

### Data availability

The data are available from the corresponding authors on a reasonable request.

### Code availability

Not applicable.

### Author contribution

Haifeng Hou conceived the study and guided the development of research and the preparation of manuscripts. Shuyu Sun, Hongzhi Liu, Zheng Guo, Qihua Guan, Yinghao Wang, Jie Wang, and Yan Qi performed the material preparation and data collection. Shuyu Sun, Hongzhi Liu, and Zheng Guo researched data, performed the statistical analyses, and wrote the manuscript. Yuxiang Yan, Youxin Wang and Jun Wen provided critical expert advice or critical review of the current manuscript. All authors read and approved the final manuscript.

## Data Availability

The data are available from the corresponding authors on a reasonable request.

## Acknowledgements

We would like to thank the participants who participated in the study for their consent and involvement for this investigation.

## Supplementary tables

Table S1 The short form of Suboptimal Health Status Questionnaire

Table S2 Results of Pearson correlation coefficients in training set (n=6,183)

Table S3 Results of extreme group method in training set (n=6,183)

Table S4 Characteristics of the cutoff points for SHSQ-SF in training set (n=6,183) Table S5 Characteristics of the cutoff points for SHSQ-SF in validation set (n=4,113)

