## Supplemental Table for "Development and Validation of a Short Form of Suboptimal Health Status Questionnaire"

**[Supplementary materials]**

**Table S1** The short form of Suboptimal Health Status Questionnaire

| **How often is it, that you (your)** | **0** | **1** | **2** | **3** | **4** |
| --- | --- | --- | --- | --- | --- |
|  | **(Never/ almost never)** | **(Occasionally)** | **(Often)** | **(Very often)** | **(Always)** |
| 1. Exhausted without physical actives significantly increasing | 🞎 | 🞎 | 🞎 | 🞎 | 🞎 |
| 2. Languid when working | 🞎 | 🞎 | 🞎 | 🞎 | 🞎 |
| 3. Muscles or joints feel stiff | 🞎 | 🞎 | 🞎 | 🞎 | 🞎 |
| 4. Have pains in shoulder/neck/waist | 🞎 | 🞎 | 🞎 | 🞎 | 🞎 |
| 5. Got out of breath while sitting still | 🞎 | 🞎 | 🞎 | 🞎 | 🞎 |
| 6. Suffered from chest congestion | 🞎 | 🞎 | 🞎 | 🞎 | 🞎 |
| 7. Got poor appetite | 🞎 | 🞎 | 🞎 | 🞎 | 🞎 |
| 8. Had trouble with impairment in short memory | 🞎 | 🞎 | 🞎 | 🞎 | 🞎 |
| 9. Could not respond quickly | 🞎 | 🞎 | 🞎 | 🞎 | 🞎 |

**Table S2** Results of Pearson correlation coefficients in training set (n=6,183)

| **Items** | **Pearson correlation coefficients** | ***P*** |
| --- | --- | --- |
| Exhausted without physical actives significantly increasing | 0.738 | <0.001 |
| Languid when working | 0.721 | <0.001 |
| Muscles or joints feel stiff | 0.761 | <0.001 |
| Have pains in shoulder/neck/waist | 0.762 | <0.001 |
| Got out of breath while sitting still | 0.738 | <0.001 |
| Suffered from chest congestion | 0.728 | <0.001 |
| Got poor appetite | 0.682 | <0.001 |
| Have trouble with impairment in short memory | 0.818 | <0.001 |
| Could not respond quickly | 0.826 | <0.001 |

**Table S3** Results of extreme group method in training set (n=6,183)

| **Items** | ***t*** | **df** | ***P*** |
| --- | --- | --- | --- |
| Exhausted without physical actives significantly increasing | 66.276 | 2702.335 | <0.001 |
| Languid when working | 68.502 | 3215.141 | <0.001 |
| Muscles or joints feel stiff | 57.621 | 1928.191 | <0.001 |
| Have pains in shoulder/neck/waist | 76.429 | 2325.766 | <0.001 |
| Got out of breath while sitting still | 46.810 | 1889.701 | <0.001 |
| Suffered from chest congestion | 48.799 | 1898.603 | <0.001 |
| Got poor appetite | 52.166 | 1974.940 | <0.001 |
| Have trouble with impairment in short memory | 78.691 | 2045.951 | <0.001 |
| Could not respond quickly | 75.265 | 1971.901 | <0.001 |

**Table S4** Characteristics of the cutoff points for SHSQ-SF in training set (n=6,183)

| **Cut-Off Point** | **Sensitivity** | **Specificity** | **Youden index** |
| --- | --- | --- | --- |
| 10.5 | 0.965 | 0.915 | 0.880 |
| 11.5 | 0.918 | 0.954 | 0.872 |
| 9.5 | 0.990 | 0.867 | 0.857 |
| 12.5 | 0.837 | 0.976 | 0.813 |
| 8.5 | 0.998 | 0.801 | 0.799 |
| 7.5 | 0.999 | 0.739 | 0.738 |
| 13.5 | 0.746 | 0.986 | 0.732 |
| 6.5 | 0.999 | 0.675 | 0.674 |
| 5.5 | 0.999 | 0.611 | 0.610 |
| 14.5 | 0.612 | 0.994 | 0.605 |
| 4.5 | 1.000 | 0.537 | 0.537 |
| 15.5 | 0.512 | 0.997 | 0.509 |
| 3.5 | 1.000 | 0.440 | 0.440 |
| 16.5 | 0.434 | 0.999 | 0.433 |
| 17.5 | 0.361 | 0.999 | 0.360 |
| 2.5 | 1.000 | 0.349 | 0.349 |
| 18.5 | 0.279 | 1.000 | 0.279 |
| 19.5 | 0.230 | 1.000 | 0.229 |
| 1.5 | 1.000 | 0.228 | 0.228 |
| 20.5 | 0.197 | 1.000 | 0.197 |
| 21.5 | 0.180 | 1.000 | 0.180 |
| 0.5 | 1.000 | 0.158 | 0.158 |
| 22.5 | 0.148 | 1.000 | 0.148 |
| 23.5 | 0.122 | 1.000 | 0.122 |
| 24.5 | 0.109 | 1.000 | 0.109 |
| 25.5 | 0.091 | 1.000 | 0.091 |
| 26.5 | 0.076 | 1.000 | 0.076 |
| 27.5 | 0.056 | 1.000 | 0.056 |
| 28.5 | 0.042 | 1.000 | 0.042 |
| 29.5 | 0.033 | 1.000 | 0.033 |
| 30.5 | 0.029 | 1.000 | 0.029 |
| 31.5 | 0.026 | 1.000 | 0.026 |
| 32.5 | 0.017 | 1.000 | 0.017 |
| 33.5 | 0.014 | 1.000 | 0.014 |
| 35.0 | 0.010 | 1.000 | 0.010 |
| -1.0 | 1.000 | 0.000 | 0.000 |
| 37.0 | 0.000 | 1.000 | 0.000 |

**Table S5** Characteristics of the cutoff points for SHSQ-SF in validation set (n=4,113)

| **Cut-Off Point** | **Sensitivity** | **Specificity** | **Youden index** |
| --- | --- | --- | --- |
| 10.5 | 0.962 | 0.874 | 0.836 |
| 11.5 | 0.905 | 0.930 | 0.834 |
| 9.5 | 0.986 | 0.814 | 0.800 |
| 12.5 | 0.825 | 0.963 | 0.788 |
| 8.5 | 0.991 | 0.722 | 0.712 |
| 13.5 | 0.700 | 0.980 | 0.679 |
| 7.5 | 0.998 | 0.638 | 0.635 |
| 6.5 | 1.000 | 0.549 | 0.549 |
| 14.5 | 0.544 | 0.992 | 0.536 |
| 5.5 | 1.000 | 0.459 | 0.459 |
| 15.5 | 0.408 | 0.997 | 0.405 |
| 4.5 | 1.000 | 0.373 | 0.373 |
| 16.5 | 0.331 | 0.999 | 0.329 |
| 3.5 | 1.000 | 0.285 | 0.285 |
| 17.5 | 0.234 | 1.000 | 0.234 |
| 2.5 | 1.000 | 0.197 | 0.197 |
| 18.5 | 0.161 | 1.000 | 0.161 |
| 19.5 | 0.118 | 1.000 | 0.118 |
| 1.5 | 1.000 | 0.109 | 0.109 |
| 20.5 | 0.090 | 1.000 | 0.090 |
| 21.5 | 0.071 | 1.000 | 0.071 |
| 0.5 | 1.000 | 0.056 | 0.056 |
| 22.5 | 0.055 | 1.000 | 0.055 |
| 23.5 | 0.043 | 1.000 | 0.043 |
| 24.5 | 0.028 | 1.000 | 0.028 |
| 25.5 | 0.022 | 1.000 | 0.022 |
| 26.5 | 0.021 | 1.000 | 0.021 |
| 27.5 | 0.013 | 1.000 | 0.013 |
| 28.5 | 0.007 | 1.000 | 0.007 |
| 29.5 | 0.005 | 1.000 | 0.005 |
| 31.0 | 0.003 | 1.000 | 0.003 |
| 34.0 | 0.002 | 1.000 | 0.002 |
| -1.0 | 1.000 | 0.000 | 0.000 |
| 37.0 | 0.000 | 1.000 | 0.000 |
